# Protocol of the study for Setting up of a system for “Medical Certification of Cause of Death” for non-institutional deaths in a selected area of a Taluk of Kolar district, Karnataka, India: feasibility and validity

**DOI:** 10.1101/2025.02.05.25321716

**Authors:** M Madhusudan, R Sukanya, K Vaitheeswaran

## Abstract

**Rationale:** The current coverage of “Medical Certification of Cause of Death” in India is only 22.5%. This is largely due to significant proportion of deaths occurring outside the hospitals (non-institutional deaths). The “cause of death” in such cases is unlikely to be certified by any doctor. The present study attempts to address this gap by developing a system of MCCD for “non-institutional deaths” in the country.

**Novelty:** This is a first of its kind study attempting to address the gap of coverage as well as reliability of MCCD for “non-institutional deaths”.

**Objectives:** To assess the feasibility of “Physician derived Cause of death” approach for deducing “cause of death” in “non-institutional deaths” in a selected area of Kolar Taluk, Karnataka and to validate this approach

**Methods:** Doctors of selected major hospitals in Kolar taluk and PHC medical officers and private practitioners of 2 selected PHC areas of the taluk would be trained in arriving at “Cause of Death” in “non-institutional deaths” using the “PhyCoD” tool. The “cause of death” deduced by this approach would be validated against the ‘gold standard’ autopsy wherever possible. The approach will also be tested for “inter-rater reliability”.

**Expected outcome:**

a. Development of a tool for physicians for deducing “Cause of Death” in “non-institutional deaths”
b. Increased coverage of MCCD for “non-institutional deaths”
c. Improved accuracy in the reporting of “cause of death” for “non-institutional deaths”
d. Reduced delay in the reporting of “cause of death” for “non-institutional deaths”

## Introduction

Reliable age and gender-wise “cause of death” statistics is required on a regular basis by Administrators, Policy Planners etc., for evidence-based decision-making with regard to identifying the priorities areas for resource allocation, monitoring of various indicators and other related activities in the area of Public Health. Keeping this in mind, the scheme of “Medical Certification of Cause of Death (MCCD)” was introduced in the country under the provisions of “Registration of Births and Deaths (RBD)” Act, 1969. However the coverage of “Medical Certification of Cause of Death (MCCD)” in India out of the total registered deaths currently is dismal at 22.5% and this low coverage has hardly improved over the last decade.^1^ This is largely attributed to significant proportion of deaths (60-70%) occurring outside hospital (deaths at home viz., deaths during sleep, sudden deaths etc., and those during travel to hospital) which are classified as “non-institutional deaths”. The “cause of death” in such cases is unlikely to be certified by any doctor.

‘Brought dead’ cases to hospital where an unnatural cause or manner of death (like accident, homicide, suicide) is suspected is sent for autopsy as per the law for determining the ‘cause of death’. ‘Brought dead’ cases where the physician does not suspect any unnatural cause are also advised to go for autopsy to determine the “cause of death”. However, it is refused by the relatives of the deceased in most of the cases. Also, there are very few hospitals in our country where medical or clinical autopsy is offered for ‘brought dead’ cases where the manner of death is ‘natural’. The physician in the hospital does/can not certify the ‘cause of death’ in such cases as he has not treated the case. Hence such deaths end up without the cause of death being documented. A large proportion of deaths at homes (viz., old age) may not even reach the hospital/doctor at all.

“The Registration of Birts and Deaths act, 1969” clearly states that for all deaths the ‘cause of death’ has to be certified by the doctor who attended to the decedent prior to his death. But it does not clarify how “cause of death” can be certified in an individual who has not received any medical care in the recent past.^1^

In recent times, most of the states have mandated the hospitals to submit the form 4 (MCCD form) for all deaths occurring in the institutions in order to complete the death registration. However, for “non-institutional deaths”, form 4A is neither insisted upon nor is there any suitable guidance provided. As a result of all these, “non-institutional deaths” (60-70% of all deaths) in India may be registered with the Local Registrar but may not have a medically certified “cause of death”.

This study attempts to develop a system to address the gap of coverage in MCCD for “non-institutional deaths”. There is some contact with medical facility just before death in 54.6% of the cases in India.^1^ Hence, it may be worthwhile to establish a system for deriving “cause of death” for “non-institutional deaths” by involving medical doctors.

This would add to the already existent methods of arriving at “underlying cause of death”: 1) MCCD for institutional deaths “(under the Civil Registration System)” and 2) “Verbal autopsy under the Sample Registration System”. VA is conducted by lay workers called SRS supervisors who collect history from the family members of the deceased using a validated tool, and the information is subsequently reviewed by physicians to derive the ‘cause of death’ [Physician Coded VA]. This two-step process is time consuming and delays assignment of CoD. Moreover, SRS is limited by the population coverage for deriving cause of mortality statistics at district level and by availability of doctors proficient to read the narratives in many regional languages.^2,3^

In this study, doctors would use a validated questionnaire to collect history of the deceased using multiple available sources of information like the relatives, sociodemographic information, medical records, investigations etc., to arrive at the CoD. Doctors would be well versed in taking history and abstracting the ‘cause of death’ information to the MCCD form. Hence this approach of “Physician derived Cause of Death”, can significantly reduce time and resources in arriving at UCoD and increase reliability of ‘cause of death’ as compared to verbal autopsy. In this context, the above study will be done to assess the feasibility of “Physician derived Cause of death (PhyCoD)” approach for deducing “cause of death” in “non-institutional deaths” in a selected area of a Taluk of “Kolar district, Karnataka” and to validate this approach.

## Materials and Methods

This would be a Cross-sectional study to be conducted in 1 government (Distt Hospital”) and 3 private hospitals (of which 1 is a Medical College) (totally 4) and 2 selected PHC areas of the Kolar Taluk of “Kolar district, Karnataka”

ICMR-NCDIR had recently conducted a baseline assessment of the MCCD system in public and private hospitals of Kolar district, Karnataka which included 1 Medical College (private), 1 District Hospital (govt), 3 Taluk Hospitals (govt), and 3 major private hospitals. In addition, Kolar is also covered by the Digital Nerve Centre of the Tata Community services that provides the linkages of primary health care through IT systems. The “Digital Nerve Centre (DiNC)” is housed in the public health system and has digitized the household data and the medical services provided through the DiNC virtual platform. Hence, Kolar District has been selected for the study.

The study would be done in 2 phases as follows (fig 2):-

**Fig 1:**
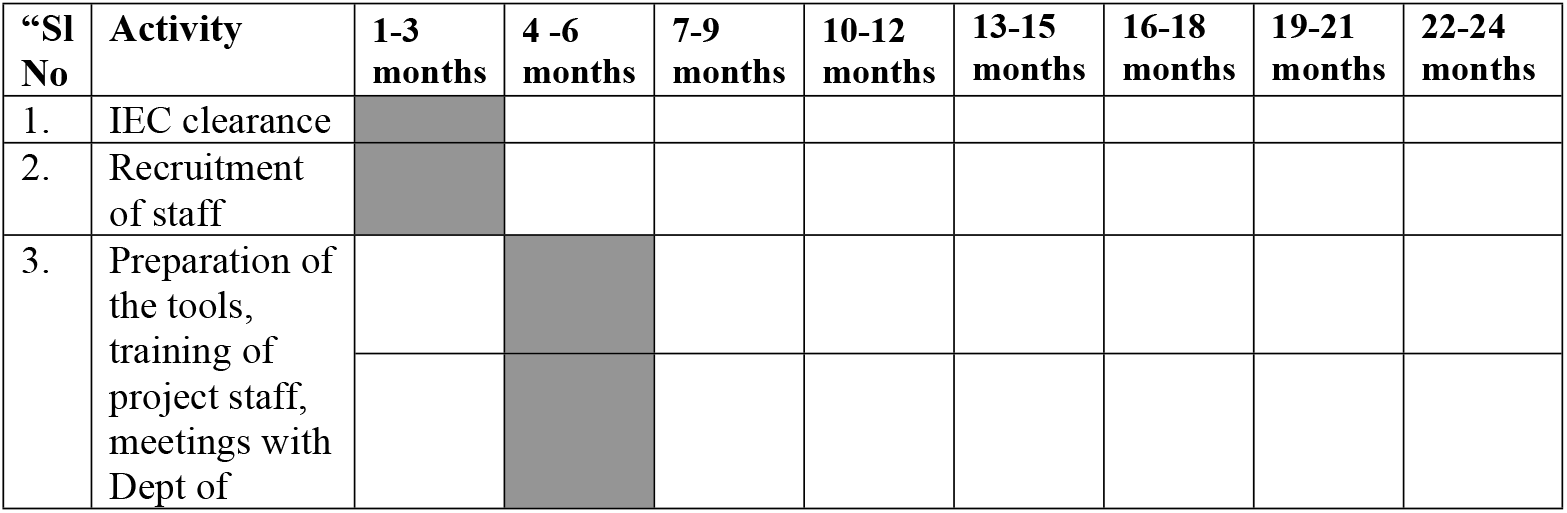
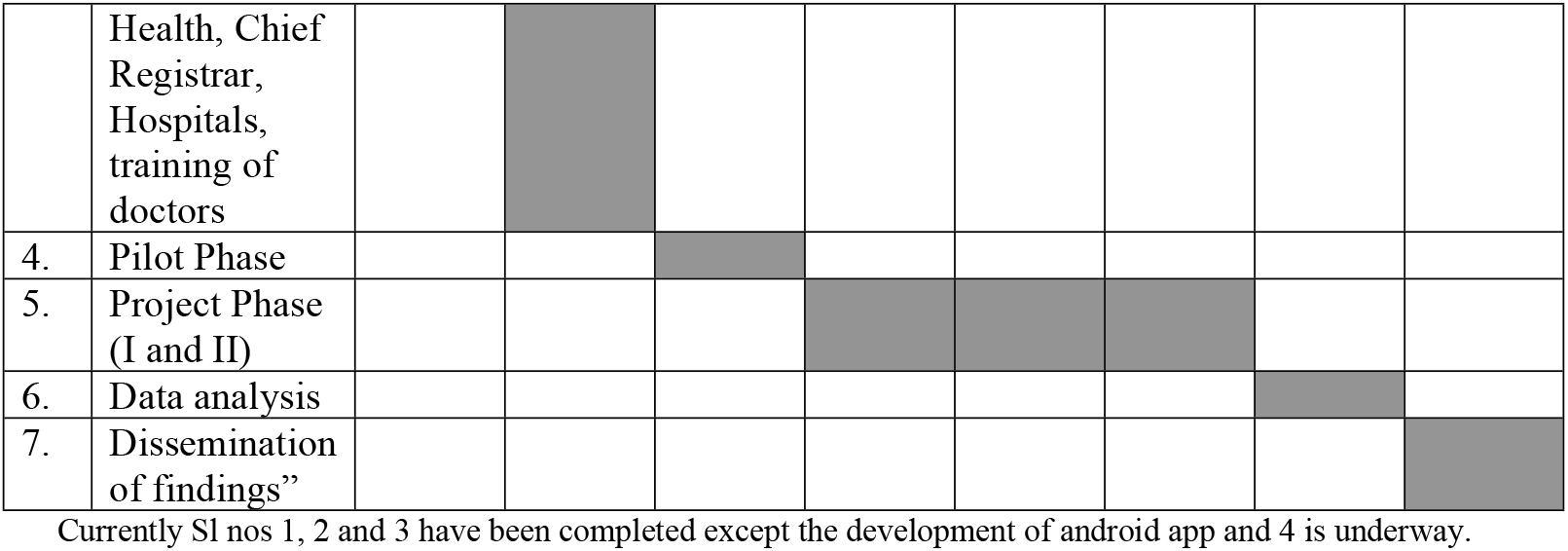
Timelines of the project

**Fig 2:**
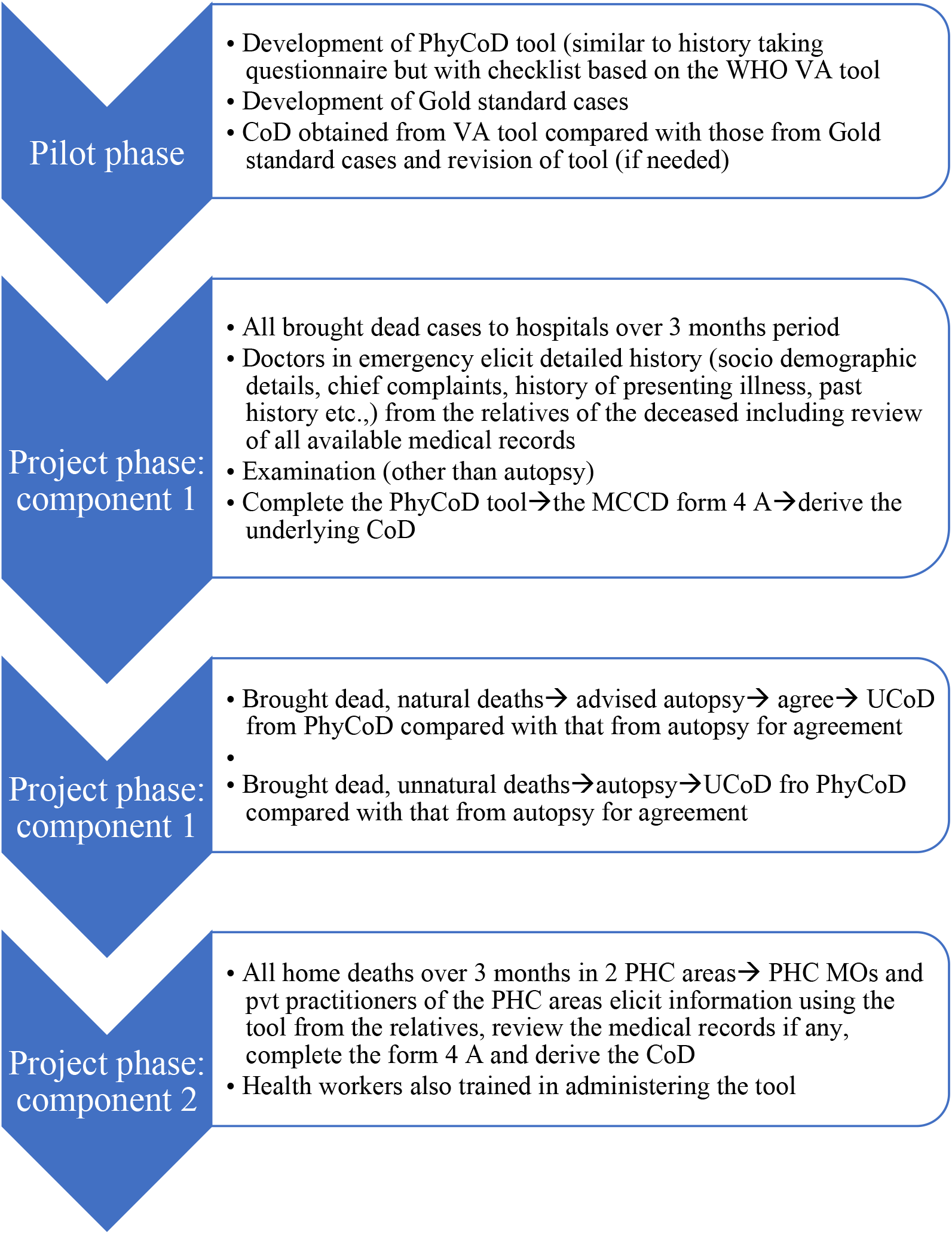
Methodology of the Pilot and project phase

I. Pilot phase: The tool for “Physician derived Cause of death (PhyCoD)” has be developed. This comprises of 4 questionnaires as follows:-
  1. Questionnaire for neonatal deaths (deaths post birth to 28 days)
  2. Questionnaire for Child deaths (deaths 29 days onwards to 14 years)
  3. Questionnaire for Maternal deaths (for female deaths in the age group of 15-49 years during pregnancy, delivery or within 42 days of delivery (including deaths during and within 42 days of abortion)
  4. Questionnaire for adult deaths (for all other deaths)

These would be similar to history taking questionnaire routinely used by doctors comprising of chief complaints, history of presenting illness, past history etc., but with a checklist based on the WHO VA tool so as to not miss any information.^4^ The chief complaints have been listed for each of these questionnaires considering the causes of death for each of these groups as per the recent MCCD report.^1^ Besides these, provision has also been made to capture others presentations if any.

During this phase, gold standard case scenarios would be developed from selected institutional deaths that have occurred over a period of last 1 year with detailed case records available. Doctors of Community Medicine department of the Medical college would be trained in MCCD and collecting information (sociodemographic, medical history) using the tool and deducing “Underlying Cause of Death” using this information. The trained doctors would administer the tool among relatives of the deceased and derive the CoD. This will be compared with the gold standard CoD from Medical records. The agreement of the physician derived CoD with Gold standard in arriving at specific “Underlying Cause of Death” and major cause group will be measured. The tool will be refined if needed and additional inputs to doctors for arriving at CoD will be accordingly provided. The tool will be finalised for deriving the “cause of death” for “non-institutional deaths” based on this.

Sample: A total of 30 cases would be taken giving proportional weightage to each of the 4 types of deaths based on the past MCCD and SRS “Cause of Death” reports of the country. Within each type of death, different types of causes of Death within the category “(viz., communicable diseases, non-communicable diseases, external cause for adult)” would be considered.

II Project phase: 4 hospitals (1 public and 3 private) that were part of the earlier baseline assessment and 2 PHC areas (1 urban and 1 rural PHC area with the largest population size) will be included. The doctors in these hospitals and Medical Officers and Private practitioners of the 2 PHC areas will be trained in MCCD and the tool to derive CoD in “non-institutional deaths”. The tool will be administered in two settings.
  1. First component would involve brought dead cases to the 4 hospitals.
  2. Second component would include all home deaths in 2 selected PHC areas of Kolar Taluk, Kolar district.

For the first component, 1 or more doctors from the each of the major departments (where deaths occur) in each of the hospitals and also Community Medicine dept (in case of Medical colleges) would be trained in MCCD and collecting information (sociodemographic, medical history) using the PhyCoD tool and deducing “Underlying Cause of Death” using this information in “Non-Institutional Deaths” and for the 2nd component, PHC Medical officers of 2 selected PHCs and leading private practitioners in these areas would be trained in the same manner. Standard Case scenarios with flowcharts for decision making so as to arrive at the most likely “cause of death” given the information (when different sets of informations are available/ unavailable) would be used in the training.

Sample: All “Non-Institutional Deaths” (deaths at home and during the travel to hospital/clinic including unnatural deaths) that occur over a 3-month period would be considered. Post-hoc power analysis will be done at the end of the study to determine the ‘power’

Component 1 (Brought dead cases to Medical College, Distt Hospital and Private hospitals): Doctors in the emergency / casualty / assigned for research study would take detailed history (Socio demographic details, chief complaints, history of presenting illness, past history etc.,) from the relatives of the deceased including review of all available medical records. They would also do the examinations *(other than autopsy)* which are relevant in arriving at the “cause of death”.^5^ Based on all these, the doctors would complete the PhyCoD tool, the MCCD form 4A and derive the underlying CoD.

For brought dead cases where no unnatural cause/manner of death is suspected, the doctors would advise the relatives to go for clinical/medical autopsy to determine the “cause of death”. In the instances where the relatives agree for clinical autopsy, the Physician derived UCoD will be compared with that from the medical autopsy report for extent of agreement. In case of medicolegal cases (where autopsy will be mandatorily conducted), the Physician derived UCoD will be compared with that from the autopsy report for agreement.

History would be collected from *one or more attendants* accompanying the deceased. They may include relatives, friends or acquaintances of the deceased who stay with the deceased and/or know the medical history of the deceased and also eyewitnesses like co-passengers, passersby, police etc., at the time of death.

Component 2: Deaths occurring over a period of 3 months at home which may or may not have sought medical attention for the illness before death in the 2 PHC areas would be considered. Information on deaths, population etc., will be obtained from “Tata Digital Nerve centre”, that is maintaining the population data and supporting clinical and community health initiatives in this district and also from the health workers of the PHC and “District Statistical Office”.

The PHC medical officers and private practitioners of the 2 PHC areas would visit the houses of the deceased after the grieving period but within 1 month of death and elicit the information using the tool from the relatives, review the medical records if any, complete the form 4 A and derive the CoD. Health workers of the PHC area will also be trained to administer the tool and collect the relevant information to complement the doctors if there are large number of home deaths. However, completion of the form 4A and deriving of the UCoD will be done by the doctor based on PhyCoD tool information.

History would be collected from one or more informants, who may include relatives, friends or acquaintances of the deceased who stay with the deceased and/or know the medical history of the deceased and also eyewitnesses at the time of death.

### Validation

i. In ‘Brought dead cases’ wherever autopsy is done, the physician derived UCoD would be compared with that in the autopsy report (gold standard) (for accuracy).
ii. In a sample of “non-institutional deaths” in the 2 PHC areas, an independent trained physician shall administer the tool to elicit detailed medical history and derive the UCoD. The agreement between the two physicians in deriving UCoD by the above-mentioned approach (i.e., history given by the relatives, and minimal medical records possessed by the relatives) would be measured (inter-rater reliability).

### Implementation strategy

The study shall be implemented by the PI,Co-PIs and the project staff from ICMR-NCDIR as per the timelines in fig 1. Preliminary meetings have been held with “Dept of Health and Family Welfare and Chief Registrar of Births and Deaths, Karnataka” to obtain permission and their collaboration in this study. Necessary directions to the hospitals and doctors for their participation in the study have been obtained from the “Dept of Health and Family Welfare, Karnataka”. Subsequently meetings have been held with the “District Health Officer”, Hospitals and Doctors to sensitize them on the project which in turn would be followed by training and implementation of the project.

Initially a PHC level meeting would be held involving the village leaders (from the local body, the religious groups, co-operative unions, etc.,), PHC MO, PHC Health workers, ASHAs, Anganwadi Workers etc. The purpose would be explained to the village leaders and their co-operation would be sought. Subsequently the households would be approached through the PHC Health workers, ASHAs, Anganwadi Workers etc., who are acquainted with the household.

### Data collection and management

A web-based tool to capture information on Neo-natal, Child, Adult and Maternal deaths is being developed using latest “Microsoft technologies and MS SQL Server”. The questionnaires shall have skip patterns, validations and in-built quality checks to capture accurate information. Access based privileges for data entry, search records and download records. The web-based application is meant for access from any location with an internet connection, and the data would be synced directly to the online database server hosted by ICMR-NCDIR.

Data would be collected using tabs by the doctors. All data collected would be stored in server at ICMR-NCDIR for subsequent retrieval and analysis.

### “Statistical analysis”

Kappa statistic would be used to measure the extent of agreement between the “physician derived UCoD” and that derived from autopsy, and also that among the doctors.

### “Ethical issues”

“Ethical clearance from the “ICMR-NCDIR Institutional Ethics Committee has been obtained (No: NCDIR/IEC/3073/2023, 21/02/2024). Written Informed consent would be obtained from the kin of the deceased in the presence of impartial witness(es). Identity of the deceased and the “cause of death” would be kept confidential. Data would be stored in password protected systems and would have access to only PI/Co investigators. Hard copies of the filled questionnaires would be stored in locked cabinets” with access only to the PI/CoPIs.

## Discussion

This study intends to address the gap of coverage as well as accuracy of MCCD for “non-institutional deaths” in the country which account for over 60% of the registered deaths in the country. The study intends to develop a tool for physicians for deducing “Cause of Death” in “non-institutional deaths”. Depending upon the outcomes of the study (feasibility and validity of the approach) the tool may be used to elicit history in “non-institutional deaths” leading to increased coverage of “non-institutional deaths” under MCCD, improved accuracy and reduced delay of the “Cause of Death” reporting for “non-institutional deaths”.

The findings of the study shall be shared with the “Office of the Registrar General of India (ORGI)”, “Chief registrar of Karnataka” and the “Dept of Health, Karnataka”. Based on the outcome of the project, the approach may be subsequently implemented in other districts and states of the country to increase the coverage of MCCD for “non-institutional deaths”.

### Challenges in implementation

There may be very limited history available in a significant proportion of cases viz.,

1. No other illnesses, sudden death in a middle aged individual
2. Old age (80+), only generalized weakness, otherwise healthy

In such cases the doctors would have to rely on investigations which may be helpful in arriving at “cause of death”. These may be refused by the kin of the deceased in a significant proportion of cases.

In a certain proportion of cases there would be limited information available in the case sheets and also from the kin of the deceased. The “cause of death” events would have to be deduced based on the available information only (for comparison of CoD events based on the case sheet against the “cause of death” events obtained by using the PhyCoD questionnaire). Also in a significant proportion of cases especially in individuals with multiple morbidities, it would be difficult to delineate the sequence of “cause of death” events from other contributory causes. These would be important challenges in validating the questionnaires.

## Data Availability

No datasets were generated or analysed during the current study. All relevant data from this study will be made available upon study completion

NA

## Acknowledgements

The authors would sincerely acknowledge the guidance and support provided by the Director, ICMR-NCDIR, Bengaluru and also administrative support provided by the Administrative staff of ICMR-NCDIR.

## Authors’ Constributions

1. Conceptualization, Drafting the manuscript
2. Drafting the manuscript, Critical review
3. Critical review

